# Evaluating Quality Adjusted Life Years in the Absence of Standard Utility Values- A Dynamic Joint Modelling Approach

**DOI:** 10.1101/2020.05.24.20112102

**Authors:** Vishal Deo, Gurprit Grover

## Abstract

Estimation of Quality Adjusted Life Years (QALYs) is pivotal towards cost-effectiveness analysis (CEA) of medical interventions. Most of the CEA studies employ multi-state decision analytic modelling approach, where fixed utility values are assigned to each disease state and total QALYs are calculated on the basis of total lengths of stay in each state.

In this paper, we have formulated a new approach to CEA by defining utility as a function of a longitudinal covariate which is significantly associated with disease progression. Association parameter between the longitudinal covariate and survival times is estimated through joint modelling of the longitudinal linear mixed effects model and the Weibull accelerated failure time survival model. Metropolis-Hastings algorithm and Monte Carlo integration are used to predict expected survival times of each censored case using the joint model. Fitted longitudinal model is further used to project values of the longitudinal covariate at all time points for each patient. Utility values calculated using these projected covariate values are used to evaluate QALYs for each patient.

Retrospective survival data of HIV/ AIDS patients undergoing treatment at the Antiretroviral Therapy centre of Ram Manohar Lohia hospital in New Delhi is used to demonstrate the implementation of the proposed methodology. A simulation exercise is also carried out to gauge the predictive capability of the joint model in projecting the values of the longitudinal covariate.

The proposed dynamic approach to calculate QALY provides a promising alternative to the popular multi-state decision analytic modelling approach, especially when the standard utility values for different stages of the concerned disease are not available.

## 1 Introduction

Calculation of Incremental Cost Effectiveness Ratio (ICER) based on gain in Quality Adjusted Life Years (QALYs) is a common technique for Cost Effectiveness Analysis (CEA) of a treatment regime. Quality of life at any stage of a disease is quantified in terms of utility values. Direct methods like visual analog scale (VAS) and standard gamble (SG) techniques, and indirect methods based on questionnaires, like EQ-5D, SF-6D, HUI2, HUI3 etc, are available to calculate standard utility values for different stages of a disease; refer [1-5]. Use of these utility values requires classification of disease progression into well defined stages. So, as a popular approach, multi-state models are used to estimate average gain in QALYs since the beginning of the treatment till the lifetime time horizon chosen for the study. Transition probabilities between different states of the disease can be estimated by fitting survival models to the transitions. Cox-Proportional Hazard (PH) models [6, 7], parametric survival models [8-11], Bayesian models [12] and flexible parametric models [13] are some of the most frequently applied models for this purpose. Transition probabilities are further used to estimate expected Total Length of Stay (TLOS) in each state till the lifetime time horizon of the study. TLOS is weighed in with respective utility value for each state and summed over all states to obtain total gain in QALY. This procedure for conducting CEA is called Markov/ multi-state Decision-Analytic Modelling; refer [14].

### Limitations imposed by multi-state decision-analytic models

Model states or disease stages are defined on the basis of one or more covariates(s) /biomarker(s) measured repeatedly at specific follow-up time points over the period of the study. These follow-up times can be at equal or unequal intervals, and are most likely to differ from patient to patient. In such cases, the exact time and path of transition from one stage to another is not known, which is likely to induce error in the estimation of transition probabilities and TLOS. Also, by compartmentalizing the progression of disease into predefined static states, we undermine the dynamics of disease progression within each state. For example, in CEA of Antiretroviral Therapy (ART) for HIV/ AIDS patients, stages are defined on the basis of CD4 cell counts measured at each visit of the patient. CD4 ≥ 500 => stage I, 350≤ CD4<499 => stage II, 200 ≤CD4<349 => stage III, and CD4< 200 => stage IV. So, while using multi-state decision-analytic modelling, five states are defined, where the fifth state is an absorbing state corresponding to death and all other states are transient and are accessible from each other. Suppose that a patient is found to be in state II at the start of ART (t=0) and in state IV at next visit (say t= 3 months). Since the information during the interval is missing, the possible transition paths could be II->III->IV or II-IV or II->I->IV and many more. This problem does not exist for diseases where the states are irreversible; however, the problem of lack of information on exact time of transitions still persists. Further, for the sake of discussion, let us define utility values for the five disease states as 1, 0.8, 0.6, 0.4 and 0 respectively. So, if a patient A has a CD4 count of 351 (stage II) at a visit and his/her next visit is after one year, utility value will be taken as 0.8 for the whole one year (i.e. 0.8 QALY gain). Whereas, if a patient B’s CD4 count is recorded as 348 (stage III) at a visit and his/her next visit is after one year, utility value will be considered as 0.6 for that one year (i.e. 0.6 QALY gain). That is, a significant difference in the calculated value of gain in QALY can exist even when, in reality, the two patients may have similar health status (ignoring other patient specific factors). Apart from this, if we assume same CD4 counts for any two patients, before and after an interval of a year, the dynamics of the changes in the CD4 count within that year may differ drastically between the patients. This problem is also not addressed by the multi-state approach.

### Issues with standard state specific utility values

While using standard utility values for specific states of a disease, calculated using direct (VAS, SG) or indirect (EQ-5D, SF-6D, HUI2, HUI3 etc.) methods, following issues may arise.

1. In multi-state decision-analytic models, utilities obtained from direct methods, i.e. through individual level preferences, may vary significantly from those obtained from indirect methods based on community level preferences, refer [4].
2. Individual level variability in utility values can exist because of certain factors and covariates like age, sex, BMI, smoking habit, income, social background, medical history etc., see for example [15].
3. Spatial variability in standard utility scores of different regions is also expected because of the differences in economic, social, political and geographic status of the target populations. Shah et. al. [16] have given a commentary on how geomarkers can be useful in assessing patient level variability in risks. To understand through an example, an HIV patient, in any stage, staying in a developed country is expected to have a better quality of life than that of a patient in the same stage in a developing or underdeveloped country. This is because, apart from having better patient-care systems at place, developed countries also experience a wider social acceptability of HIV patients. However, on account of unavailability of region/ country specific standard utility values, researchers have time and again used standard utility values calculated for other regions/ countries with drastically different socio-economic and demographic conditions to conduct CEA. For example, while calculating gain in QALY for HIV/ AIDS patients undergoing ART in India, Bender et. al. [17] have obtained utility values from the studies in United States.
4. Using questionnaire based techniques to obtain utility values can be challenging for certain diseases when, because of lack of social acceptability and fear of discrimination, patients are hesitant in revealing their identities.
5. With fast paced changes in the patient care facilities and socio-economic standard of life, ideally, utility values should be updated regularly. This would consume a lot of financial and temporal resources.

The objective of this paper is to present a non multi-state approach to calculate QALY, using joint-modelling of longitudinal and survival data, and a proxy utility function. It is a more dynamic alternative to the multi-state decision analytic model approach. The idea is to define a function which calculates utility for each patient at each discrete time point (as per the unit of measurement of time considered in the study), say at each month, since the start of the treatment till death or till the lifetime time horizon of the study (for censored cases). This proxy utility function can be defined as a function of time-dependent covariates having statistically significant impact on hazard or survival time. Thus, this approach captures the continuous changes in quality of life of each patient separately, taking care of the patient level variability in utility values. Further, it traces the entire trajectory of disease progression instead of just monitoring transitions between few discrete states. Since, we are considering diseases where longitudinal repeated measures on some covariate(s)/ biomarker(s) are available along with the survival data, joint modelling of longitudinal and survival data is proposed for estimating the association of longitudinal variable(s) with hazard or with survival times. The estimated association parameter(s) is/are used to define weight(s) of the corresponding covariate(s) in the proxy utility function.

Proposed methodologies for the construction of proxy utility function, and for calculating QALY, are discussed thoroughly in the next section. In section 3, implementation of the proposed methodology is demonstrated on a retrospective data of HIV/AIDS patients undergoing ART at Ram Manohar Lohia hospital in New Delhi. A simulation exercise aimed at assessing predictive capability of the joint model is presented in section 4. Discussion on the results and concluding remarks are presented in section 5.

## 2 Methodology

Steps involved in the proposed methodology are listed below in the order of execution.

### 2.1 Joint Modelling of Longitudinal and Survival Data

In most of the studies related to chronic diseases, longitudinal data of patients comprises of repeated-measures on one or more time dependent covariates, in addition to the time-to-event data on the event of interest. For example, in HIV studies, patients’ follow-up data includes time till the occurrence of the event of interest (death or development of AIDS) and repeated measurements on the time-dependent biomarker (or covariate) CD4 lymphocyte count. Separate modelling of the longitudinal process and the time-to-event process fails in establishing association between the longitudinal progression of the covariate and the occurrence of the event. If our interest lies in assessing the impact of time-dependent covariate(s) on survival time, joint modelling serves as a better approach. Separate modelling does not take into account the fact that longitudinal measures on the time-dependent covariate are interrupted by occurrence of the event, thus introducing bias in the estimation; see [18]. Joint modelling approach overcomes this problem by maximizing the log-likelihood corresponding to the joint distribution of the longitudinal and time-to-event outcomes. Lucid descriptions of this approach can be found in the works of Tsiatis and Davidian [19], Yu et al. [20] and Rizopoulos [21].

#### Joint Model specification

Notations used in the specification of sub-models.

*n*: number of cases (patients).

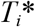: True observed event time for i-th patient.

*C_i_*: Censoring time for i-th patient.

*T_i_*: event time for the i-th patient, where 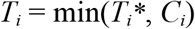

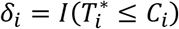; is the event indicator which takes value 1 when the event has occurred, and 0 when observation is censored.

*y_i_*(*t*): value of the observed/ measured longitudinal outcome for patient *i* at time *t*.

*t_ij_*: *j*-*th* occasion (time point) at which longitudinal response variable is observed for *i*-th patient; where (*j* = 1, 2,…, *n_i_*), i.e. number of times, and the time points at which, longitudinal responses are recorded for a patient can differ from patient to patient.

*y_ij_*: value of the longitudinal outcome for *i*-th patient at *t_ij_*.

*m_i_*(*t*): true (unobserved) value of the longitudinal outcome for *i*-th patient at time *t*. It is different from *y_i_*(*t*) as it is assumed to be free of any measurement error which is present in *y_i_*(*t*).

#### Longitudinal sub-model: Linear mixed effects model

Using the notations explained above, linear mixed effects model for i-th subject can be defined as follows; from [21].

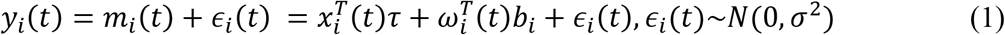

Where, *τ* denotes the vector of the unknown fixed effects parameters, *b_i_* denotes a vector of random effects, *x_i_*(*t*) and *ω_i_*(*t*) denote row vectors of the design matrices for the fixed and random effects respectively, and *∊_i_*(*t*) denotes the measurement error term which follows Normal distribution and is assumed to be independent of *b_i_*. As a standard choice, the random effects are assumed to follow multinomial normal distribution.

#### Survival sub-model: Weibull Accelerated Failure Time (AFT) Model

Again, using the notations as mentioned above, the survival sub-model can be defined as follows,

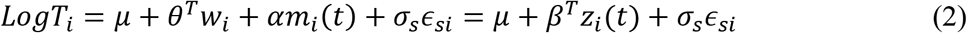

Where, *w_i_* is a vector of baseline covariates and *θ* is a vector of coefficients corresponding to these baseline covariates. *α* measures the effect of the longitudinal outcome variable, *m_i_*(*t*), on the log of survival time and is known as the association parameter. The name ‘association parameter’ emphasizes on the fact that this parameter measures the extent of association between the course of the longitudinal outcome variable and the length of the survival time. Association parameter helps in structural implementation of the idea of joint modelling of the longitudinal and survival processes. *σ_s_* is a scale parameter and *∊_si_* are independently and identically distributed random error terms following a Gumbel distribution, also known as extreme value distribution, see [22]. Under this assumption, survival time, *T_i_*, follows Weibull distribution with probability distribution function (pdf), and survival function given in the equations (3) and (4) respectively.

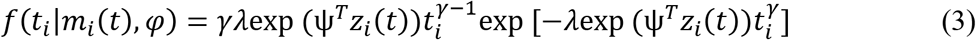

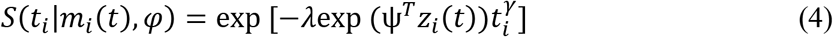

Where, 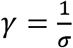, 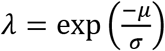, and 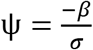 Parameters of both sub-models are estimated jointly by maximizing the joint likelihood function of the longitudinal and survival components. In our previous work on using joint modelling for utility calculations, we have used Cox proportional hazards (PH) model as the survival sub-model [23]. Weibull AFT can be considered as an improvement on the Cox PH model, as in the former case, instead of choosing lifetime time horizon of the study for censored cases, we can predict the lifetime for each patient. Thus, carrying out sensitivity analysis for the choice of lifetime time horizon, as suggested by many authors (see for example [24]), will not be required in the case of joint modelling approach with a Weibull AFT sub-model.

### 2.2 Estimating Conditional Expected Survival Time for Each Censored Case

For each censored case, conditional expectation of survival time, given that the patient has survived till time *C_i_*, is estimated and taken as patient level lifetime time horizon for calculating QALY. Conditional probabilities, *P*(*T_i_=t_i_*|*T_i_ > C_i_*), are calculated using estimated fixed effect parameters and empirical Bayes realizations of the vector of random effects *b_i_* from their posterior distribution. Metropolis-Hastings (MH) algorithm with independent proposals from a multivariate normal distribution, with mean centered at current estimate of *b_i_* and variance-covariance matrix estimated from joint modelling, is used to generate *L* empirical Bayes realizations on the vector of random effects *b_i_*, where *L* is a large number, say 100 or 1000.

Conditional expected survival times for censored cases are calculated at each of these *L* realizations of *b_i_*, using Monte Carlo integration technique. Median of these *L* values calculated for a censored case is taken as the estimate of the conditional expected survival time for that case.

### 2.3 Projection of CD4 Cell Count

Using the fitted longitudinal sub-model, CD4 count is projected for all patients at each time unit, after registration into ART till the predicted conditional expected survival time (for censored cases) or till the time of death (observed cases). At each time point, *L* empirical Bayes realizations are generated on the vector of random effects using MH algorithm. These realized values of *b_i_* and the estimates of fixed effects are used in the model defined in (1) to calculate *L* values of CD4 count at each time point. Mean of these *L* values at a time is taken as the projected/ predicted value of true CD4 count at that time.

### 2.4 Proxy Utility Function

The proposed proxy utility function is defined as a function of relative changes in repeated measures on the longitudinal outcome *y_i_*(*t*). Utility function for *i*-th patient at time *t*, *U_i_*(*t*), is defined as follows.

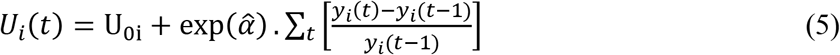

Where *U_0i_* is the base utility value of *i*-th patient at the start of the study and is defined as, 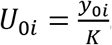. Here, *y_0i_* is the baseline value of the longitudinal covariate and *K* is the cut-off value or reference value of the longitudinal covariate beyond which it is considered to be in medically normal range. 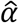, the estimate of association parameter in the survival sub-model, represents the direct impact of changes in longitudinal covariate on the log of survival time and hence, 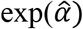 qualifies to act as a weight for determining change in the utility value as a result of relative change in the value of longitudinal covariate. Thus, the first term in the RHS of equation (5) is the baseline utility value and the subsequent terms, included in the summation, calculate the change in utility values at each time point.

This utility function is proxy in the sense that it is not based on patients’ self evaluation of their quality of life. However, it is defined as a function of a longitudinal covariate/ biomarker which is significantly associated with disease progression and acts as a proxy indicator of the quality of life of patients. It should be noted that even in the case of multi-state decision-analytic modelling approach, the longitudinal covariate has a vital role in defining utility values. For example, in the case of HIV/ AIDS patients, different states are defined on the basis of the longitudinal covariate CD4 count, and further, each state is assigned a fixed utility value. That is, in the case of multi-state model, CD4 count decides state and the state, in turn, implies a predetermined utility value. While in our approach, longitudinal progression of CD4 count directly determines utility value at any time. Projected values of CD4 count are used in the utility function defined in (5) to calculate utility values of each patient at each time unit, after registration till death or till the predicted survival time.

### 2.5 Calculation of QALY

Since, utility values are calculated at intervals of one time unit, total gain in QALY of *i*-th patient at a discount rate of *d*% per annum converted per unit time can be stated as follows.

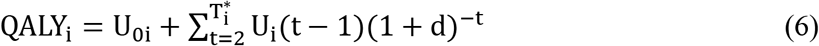

## 3 Implementation of the Proposed Methodology

As a demonstration, the methodology proposed in section 2 has been employed to calculate QALY for HIV/ AIDS patients undergoing treatment at the ART center of Ram Manohar Lohia hospital in New Delhi. Retrospective follow-up data on 3465 patients, enrolled for ART treatment during the period April 2004 to November 2009, and followed till December 2010, is obtained from register records of the hospital. All records are in terms of patient id and in no way, privacy or anonymity of patients is compromised. Cases with incomplete or missing information on factors and covariates which are included in the model, like age, sex, mode of transmission (MOT), smoking habit, drinking habit, haemoglobin, baseline weight etc., have been excluded from the analysis. Only adults of age 18 and above are retained in the study. Cases with no subsequent visit after the date of registration into ART are also not considered for the analysis. After applying all exclusion criteria, only 1520 cases qualified to be included in the study. Dataset contains information on status (dead or alive), time of death, age at the time of registration (baseline age), sex, smoking habit, alcohol consumption, baseline haemoglobin, mode of transmission of HIV (MOT), time of subsequent visits, CD4 count at each visit, and weight measured at each visit. Use of this data complies with the ethical guidelines defined in the National Ethical Guidelines for Biomedical and Health Research-2017 for administrative and secondary data. List of variables included in the longitudinal and survival sub-models are shown in Table 1. Kaplan-Meier curve of the survival data is furnished in Graph 1.

**Graph 1:**
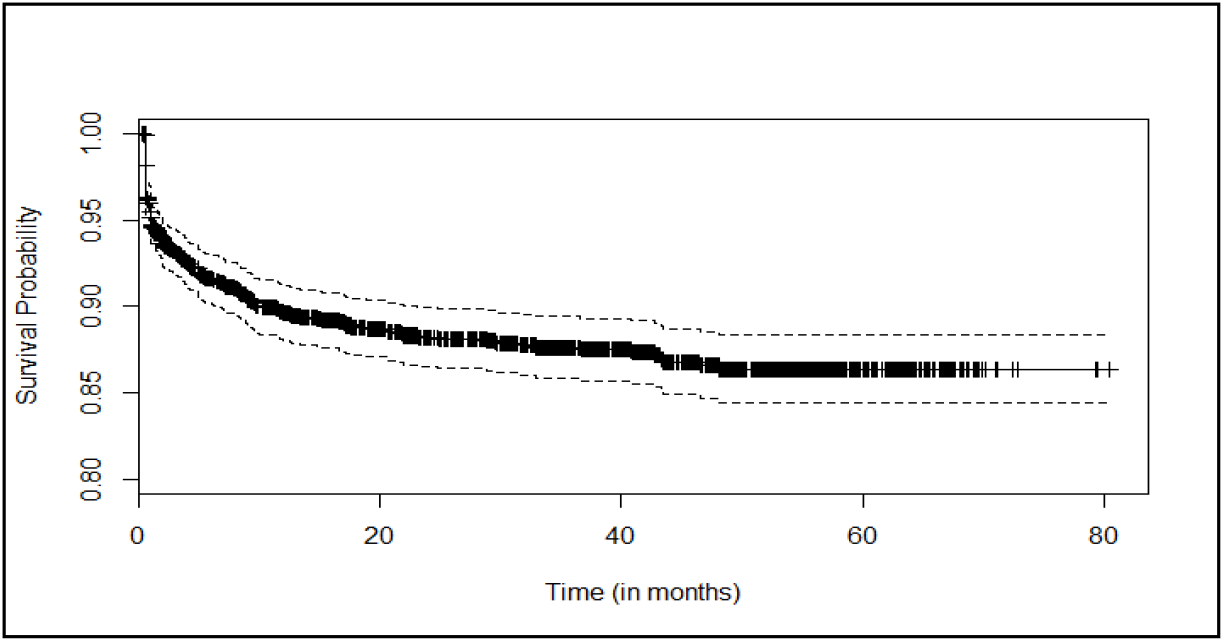
Kaplan-Meier curve for survival of HIV/ AIDS patients

**Table 1:** Description of factors and covariates included in the models

| Sl. No. | Variable name | Label | Levels (if a factor) |
| --- | --- | --- | --- |
| 1. | obstime | Time of observation in months calculated from the date of registration (a time dependent covariate) | NA |
| 2. | SEX | A factor variable with two levels, Male and Female. | 0= Female<br>1= Male |
| 3. | HB | Haemoglobin level observed at baseline | NA |
| 4. | D1_MOT | Dummy variable for mode of transmission of HIV | 1=sexually transmitted<br>0= other modes |
| 5. | D1_smoking | Dummy variable for current status of smoking | 1=currently smoke<br>0= otherwise |
| 6. | D2_smoking | Dummy variable for past status of smoking | 1=smoked in past<br>0= otherwise |
| 7. | D_alcohol | Dummy variable for status of alcohol consumption | 1= Yes<br>0= No |
| 8. | AGE | Age at baseline | NA |
| 9. | wt | Weight measured at baseline | NA |

A linear mixed effects longitudinal sub-model with response variable as CD4 count and a Weibull AFT survival sub-model are jointly fitted using the R package “JM” with method “Weibull-AFT-GH”; see [21]. Estimated models are presented in Table 2. Estimates of variance components of the longitudinal sub-model are provided in Table 3.

**Table 2:** Results of jointly fitted models

| Factor/ Covariate | Longitudinal Sub-model<br>Response : CD4 count |  | Survival sub-model<br>Response: Log T |  |
| --- | --- | --- | --- | --- |
|  | Coefficient Estimates | p-value | Coefficient estimates | p-value |
| (Intercept) | 104.4355 | **<0.0001\*\*\*** | -2.9840 | **0.0009\*\*\*** |
| obstime | 12.6425 | **<0.0001\*\*\*** | ----- | ----- |
| SEX (male) | -31.5456 | **<0.0001\*\*\*** | -0.3807 | 0.2880 |
| HB | 11.8862 | **<0.0001\*\*\*** | 0.2876 | **<0.0001\*\*\*** |
| D1_MOT (sexual) | -15.3432 | **0.0646\*** | 0.3767 | 0.2294 |
| D1_smoking (currently) | 7.6939 | 0.4204 | -0.7175 | **0.0981\*** |
| D2_smoking (past) | -4.3852 | 0.5768 | -0.7268 | **0.0280\*\*** |
| D_alcohol (yes) | -4.0831 | 0.5807 | -0.0764 | 0.8096 |
| AGE | -1.1472 | **0.0003\*\*\*** | -0.0192 | 0.1361 |
| wt | ----- |  | 0.0926 | **<0.0001\*\*\*** |
| Association | ----- |  | 0.0126 | **<0.0001\*\*\*** |
| Log(shape) |  |  | -0.3201 | **0.0003\*\*\*** |

\*\*\* Significant at 1% level of significance
\*\* Significant at 5% level of significance
\* Significant at 10% level of significance

**Table 3:**
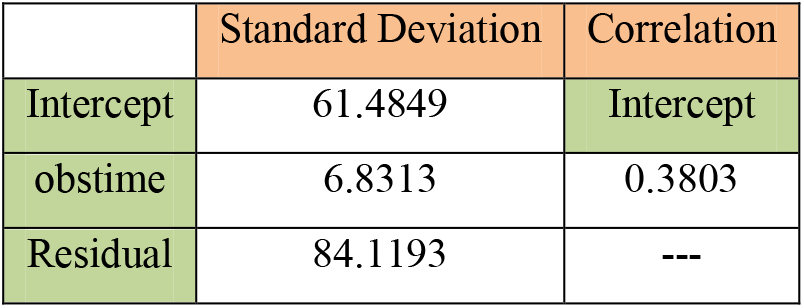
Estimates of variance components of the longitudinal sub-model

|  | Standard Deviation | Correlation |
| --- | --- | --- |
| Intercept | 61.4849 | Intercept |
| obstime | 6.8313 | 0.3803 |
| Residual | 84.1193 | --- |

Prior distribution of the random effects, *b_i_*, in the longitudinal sub-model, is taken as a multivariate normal distribution with a zero mean vector and variance-covariance matrix, *D*, obtained from the estimates of variance components of the joint modelling given in Table 3. For any censored case i, unnormalised posterior pdf of *b_i_* can be written as follows.

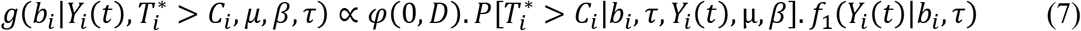

In the expression on the RHS of (7), the first term is the prior pdf of the random effects vector *b_i_*, the second term is the survival function at time *C_i_* given the values of *b_i_*, and the third term is the pdf of the response variable, CD4 count, given the values of the vector *b_i_* in the longitudinal sub-model. Given *b_i_, Y_i_*(*t*) also follows normal distribution with mean *m_i_*(*t*) and variance as estimated from joint modelling. MH algorithm with independent proposals from multivariate normal distribution, with mean centered at current estimate of *b_i_* and variance-covariance matrix estimated from joint modelling, is used to generate empirical Bayes realizations on *b_i_* from its posterior distribution. It should be noted that MH algorithm does not require the posterior distribution to be normalized. Further, for each uncensored case, following steps are followed.

1. For *i*-th patient, using MH algorithm, 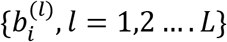 values of random effect vector are realized from the posterior distribution of *bi*.
2. At each 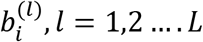, Monte Carlo integration is applied to find the conditional expected survival time for *i*-th patient given by the expression,

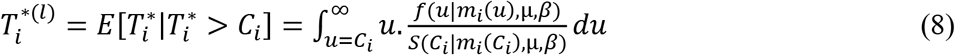
3. Median of these *L* values, 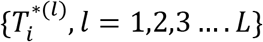, is taken as the estimate of the conditional expected survival time for the *i*-th patient, say 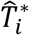.
4. For both censored and uncensored cases, at each month, 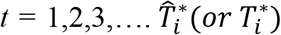, *L* realizations are generated on the vector *b_i_* and subsequently, *L* estimates of CD4 count are calculated at each month, say 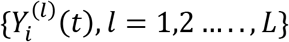. Average of these *L* values is taken as the final estimated or predicted value of CD4 count at time *t* for *i*-th patient.
5. To define the proxy utility function, we have taken *K*=800. This cut off value of CD4 count for a healthy person in India is taken on an average after examining the findings of Rungta et al. [25], Shahapur et al. [26], Ramalingam [27], and Uppal et al. [28]. CD4 counts estimated/ predicted in step 4 are used in the equation (5) to calculate utility values of each patient at each month, since registration till death or till the predicted survival time. At a standard discount rate of *d* = 3% per annum, total QALY gained by each patient is calculated using equation (6).

We have implemented these five steps in R through self-developed algorithms and programming codes. Summary of concluding results obtained from these steps is provided in Table 4.

**Table 4:** Summary of results on predicted survival times and QALY

| Outcome (in years) | Minimum | Maximum | Mean | Median |
| --- | --- | --- | --- | --- |
| Predicted Survival Time (since registration; censored cases) | 0.039 | 82 | 17.125 | 8.106 |
| Observed and predicted survival times (since registration; all cases) | 0.025 | 82 | 15.204 | 6.586 |
| QALY (discounted at 3% per annum) | 0.0002 | 28.52 | 7.46 | 4.85 |

## 4 Simulation Exercise

Using the joint model formulation discussed in section 2.1, we have simulated an individual patient level data on survival of 1000 HIV/ AIDS patients assumed to be followed for 60 months since registration into ART. Corresponding values of different baseline factors and covariates, and monthly values of the longitudinal covariate, CD4 count, are jointly simulated along-with the survival times. The R package ‘simsurv’ is used for implementation of the simulation scheme; for the simulation algorithm; refer [29]. We fit joint model to the simulated data assuming that the longitudinal observations on CD4 count are available only till 30 months from the time of registration. That is, we use only partial information on the longitudinal covariate to train the joint model. Models are fitted for the following two cases. Case 1-five observations on CD4 count (one at baseline and rest at four randomly selected time points between 1^st^ and 30^th^ month/ time-to-death) are available for each patient, and Case 2-ten observations on CD4 count (one at baseline and rest at nine randomly selected time points between 1^st^ and 30^th^ month/ time-to-death) are available for each patient. These two cases are considered to assess the sensitivity of the predictions on the number of observations available on the longitudinal covariate. Based on the results of the fitted models, the implementation steps discussed in section 3 are used to predict/ estimate CD4 counts for each month since the time of registration till the 60^th^ month (censored cases) or till the time-to-death (observed cases). Since the proposed definition of utility function depends on the longitudinal CD4 counts, credibility of the proposed method relies largely on the accuracy of prediction of the unobserved CD4 counts. So, the objective of this section is to appraise the capability of the models fitted to the training data to predict the longitudinal measurements on CD4 count beyond the observed points. Comparison between the true and predicted values of the CD4 counts are presented through graphs and the associated error is measured as Root Mean Squared Error (RMSE). For both cases, predicted values of CD4 counts, corresponding 95% credible intervals, and true values of CD4 counts for four randomly chosen censored patients and four randomly chosen observed patients, are provided in the Graphs 2-5. For the same patients, RMSE values for all cases are provided in Table 5.

**Graph 2:**
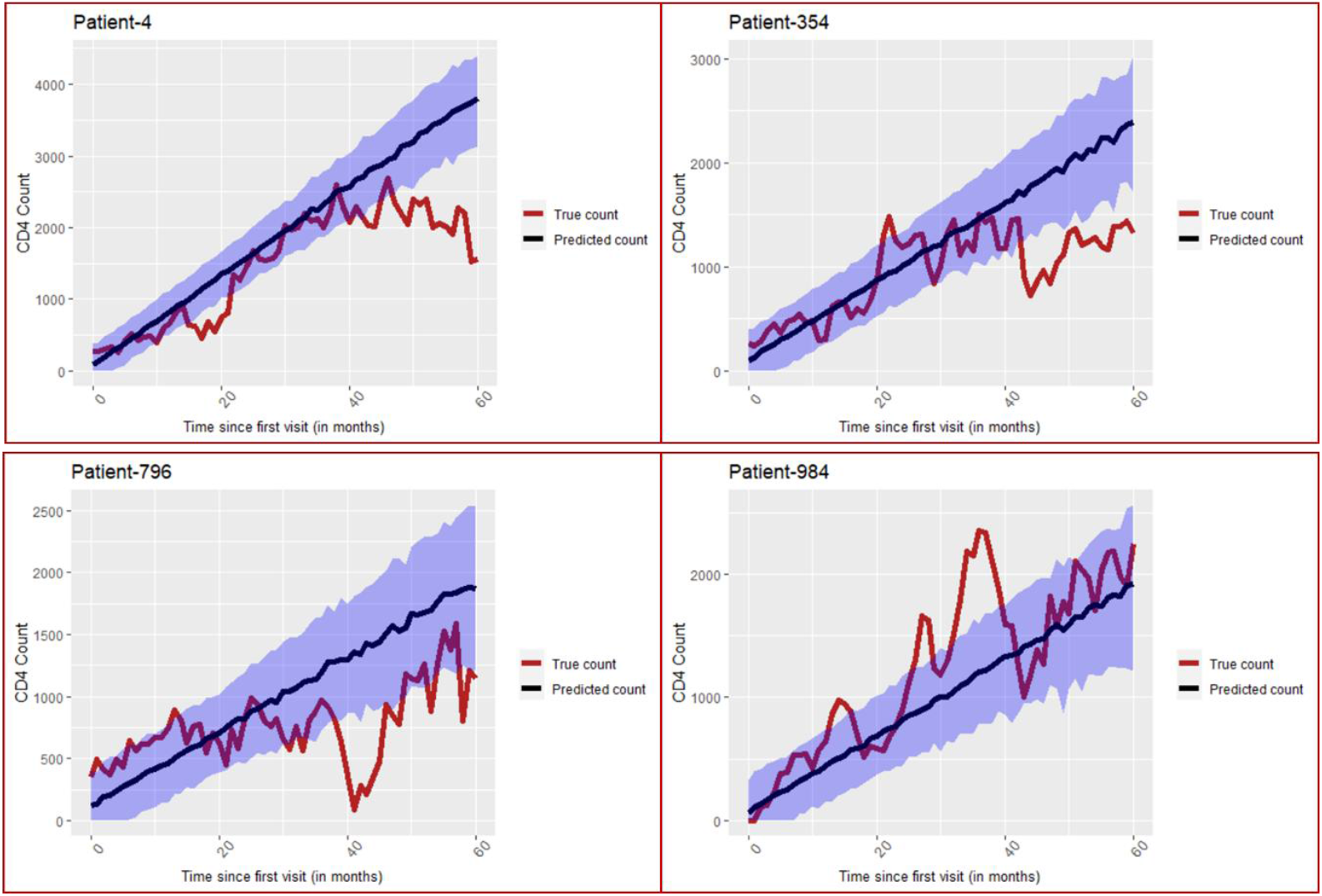
Simulation exercise-Predicted and true CD4 values for censored patients from Case 1. The blue region depicts the 95% credible interval of the predicted CD4 counts.

**Graph 3:**
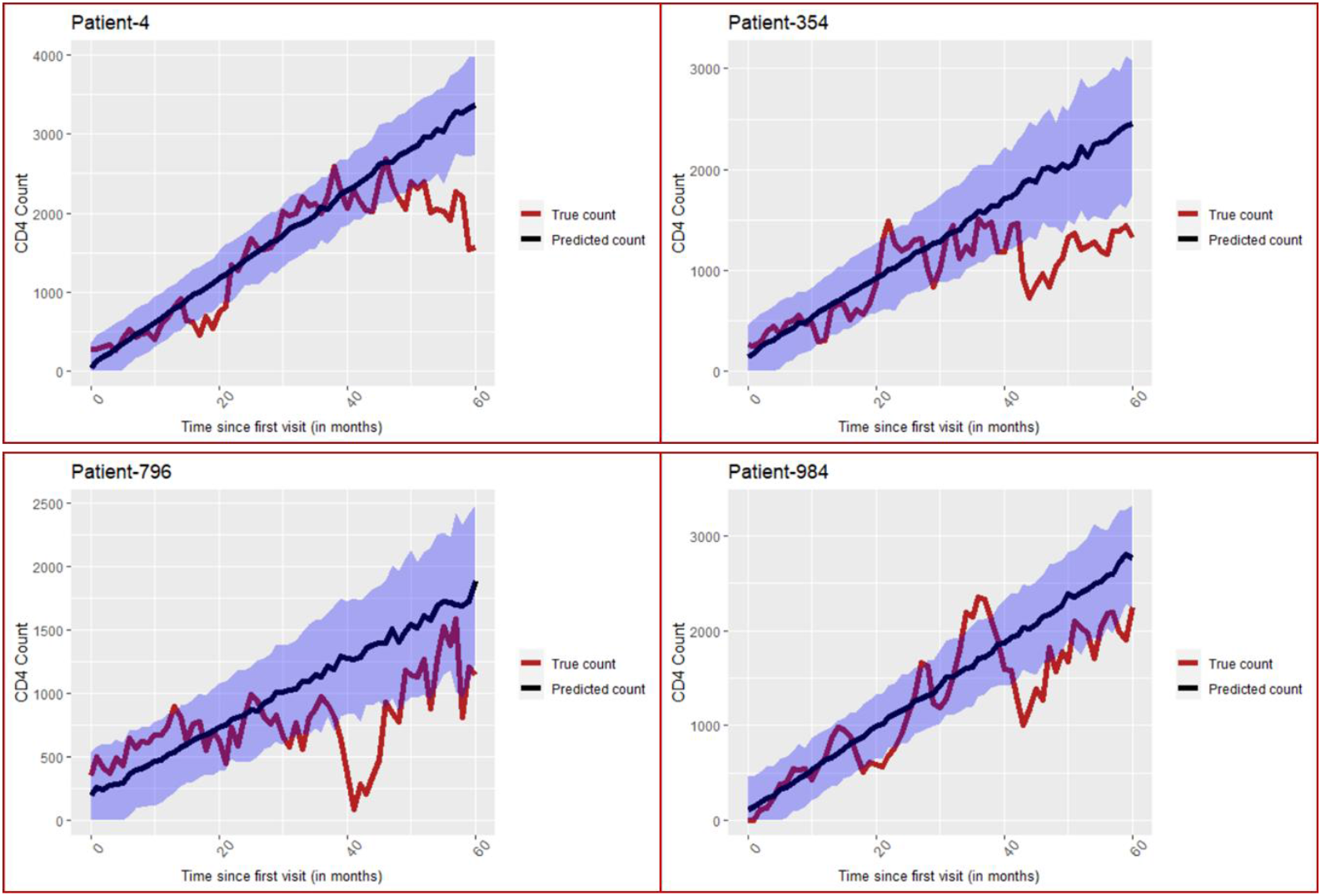
Simulation exercise-Predicted and true CD4 values for censored patients from Case 2. The blue region depicts the 95% credible interval of the predicted CD4 counts.

**Graph 4:**
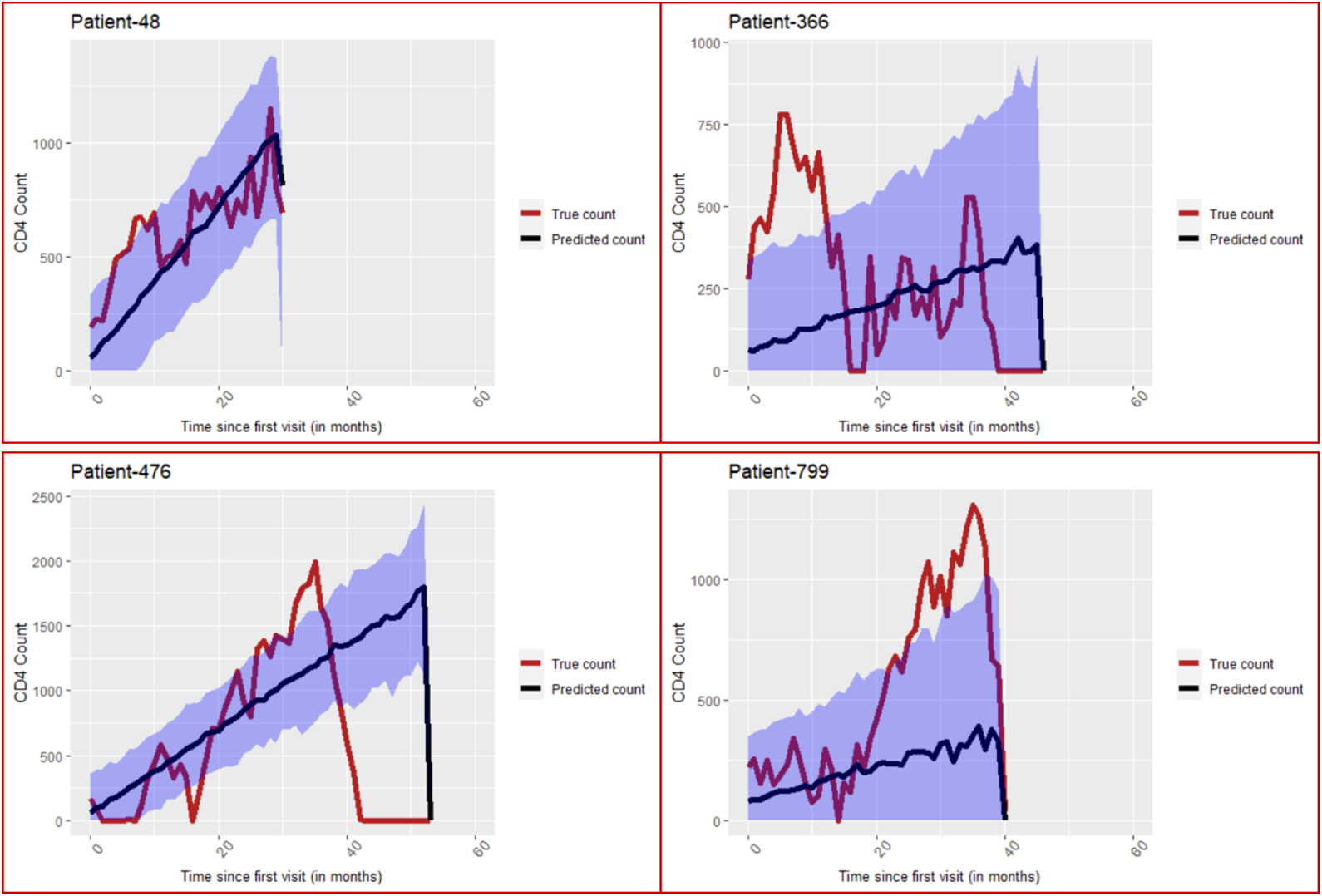
Simulation exercise-Predicted and true CD4 values for observed patients from Case 1. The blue region depicts the 95% credible interval of the predicted CD4 counts.

**Graph 5:**
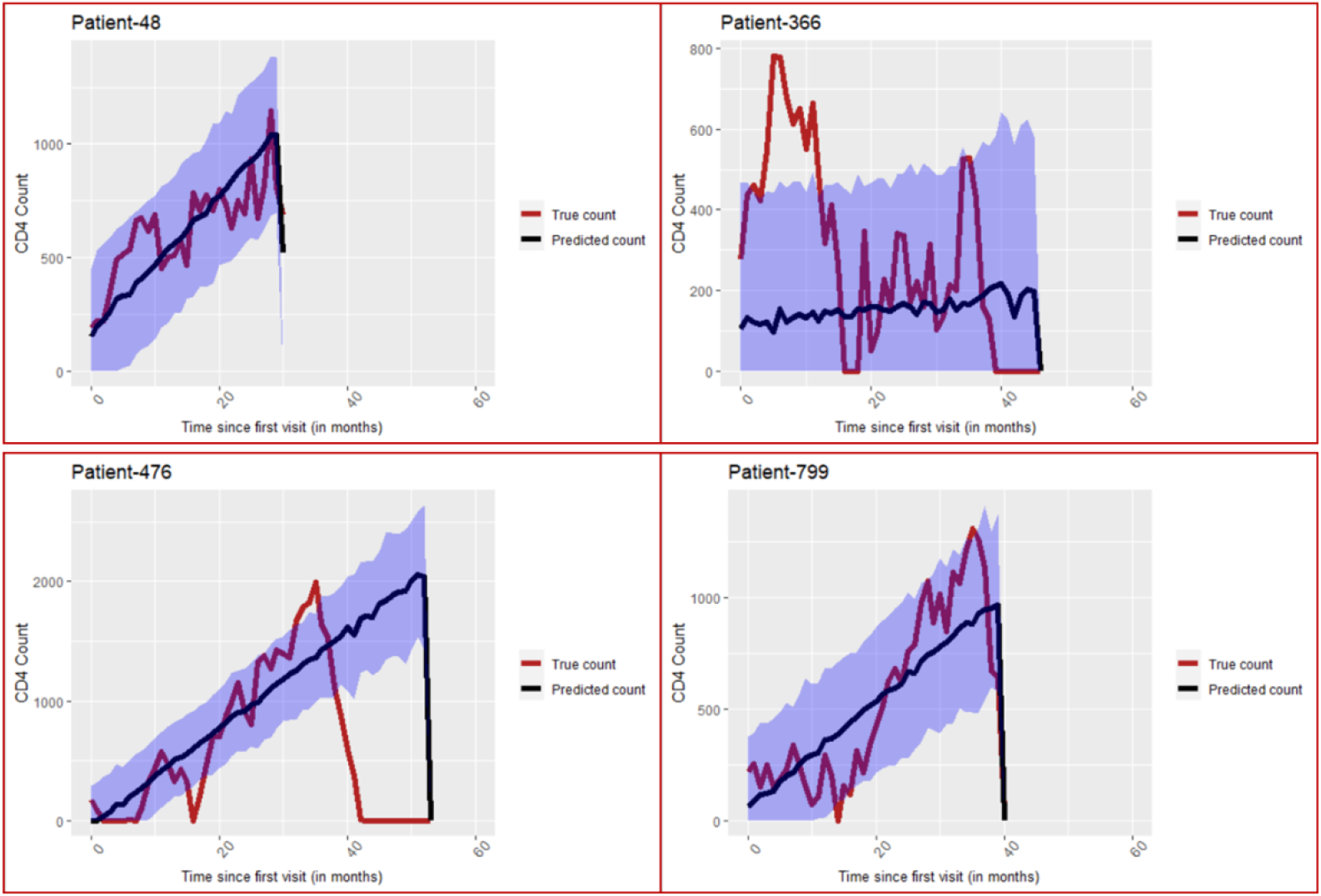
Simulation exercise-Predicted and true CD4 values for observed patients from Case 2. The blue region depicts the 95% credible interval of the predicted CD4 counts.

**Table 5:** RMSE values for all four cases

| Censored Patients |  |  |  |  |
| --- | --- | --- | --- | --- |
| Case | Patient-4 | Patient-354 | Patient-796 | Patient-984 |
| Case 1 | 743.75 | 524.66 | 510.55 | 413.24 |
| Case 2 | 536.12 | 514.34 | 459.79 | 412.09 |
| Observed Patients |  |  |  |  |
| Case | Patient-48 | Patient-366 | Patient-476 | Patient-799 |
| Case 1 | 190.64 | 309.1 | 789.38 | 449.1 |
| Case 2 | 150.21 | 279.1 | 704.19 | 204.23 |

## 5 Discussion and Conclusion

### Implementation on the retrospective ART data

Mean QALY gained by HIV/ AIDS patients after getting enrolled into ART is 7.46 years (discounted at 3% per annum). Bender et al. [17], while performing CEA for different combinations of drugs in ART in India, found that the average gain in QALY, discounted at 3% per annum, ranged from 115.5 months (9.625 years) to 125.8 months (10.48 years). One of the possible reasons for lower estimate of mean QALY in our study, as compared to that of Bender et al., is that a large proportion of patients included in our study were registered for ART at the AIDS stage, i.e. at the stage when CD4 count was less than 200. The mean baseline CD4 count of the subjects included in our study is 146.0145 (SD=104), while the mean baseline CD4 count reported by Bender et al. is 318 (SD=291). Patients with higher CD4 count at the baseline are expected to have higher survival probabilities. This is apparent from the statistically significant positive value of the association parameter estimated through joint modelling, see Table 2. Further, it is also important to note that the work of Bender et al. exhibits one of the problems discussed in our introduction section regarding the choice of utility values. They have used stage-specific utility values from published reports of US based studies to calculate QALY of patients in India. By using proxy utility function, we have eliminated the bias arising out of unaccounted regional variability in the quality of life.

### Simulation exercise

As can be seen from the Graphs 2-5, the true values of monthly observed CD4 count in the simulated data fluctuate significantly over time. Any real life data on HIV/ AIDS patients can be rarely expected to show such magnitude of fluctuations. That is, the simulated data presents a big challenge for the joint model to produce reliable predictions. Barring few cases, the predicted values of the CD4 counts can be seen to be centred around the mean trend of progression of the true CD4 counts. Predictions based on the second case are closer to the actual values, and subsequently the RMSE values for Case 2 are consistently lower than those for Case 1. The 95% credible intervals of predictions from Case 2 are narrower and better aligned with the true progression of CD4 counts as compared to those from Case 1. These findings corroborate that models fitted using more repeated measures on the longitudinal covariate of each patient show improved prediction capability. Further, predictions for observed cases show relatively much lower RMSE than those for the censored cases.

### Conclusion

The proposed method allows us to carry out CEA without categorizing disease progression into discrete states. It succeeds in gauging the dynamic effects of continuous (or short term) changes in longitudinal covariate on the utility values of the patients. Unlike multi-state decision-analytic modelling approach, this approach facilitates calculation of time-varying utility measures and hence, the total gain in QALY, of each patient. This helps to accommodate for individual level variability in the trajectory of disease progression and the quality of life. This method will be categorically useful in situations where standard utility values for the concerned disease or for the concerned population (or region of study) are not available. Even in many big economies, like India, standard utility values for most of the chronic diseases are still not available.

### Limitations

We have focussed on introducing the proposed methodology and demonstrating its implementation techniques. The simulation exercise included in this paper gives some idea about the prediction capability of the proposed model, but it is not sufficient to conclusively establish its predictive power, and hence the accuracy of utility calculations. This issue of predictive power forms the central objective of our next research work which is currently under progress.

## Data Availability

Data has been obtained with proper permission from competent authorities from the registered records of ART center of Ram Manohar Lohia hospital in New Delhi. To ensure patient confidentiality, the data is not available in public space.

